# Validation and Sensitivity Analysis of the COVID-19 Transmission Model Simulating Counterfactual Infections in Japan

**DOI:** 10.1101/2025.02.08.25321916

**Authors:** Hideki Kakeya, Yoshitaka Umeno

## Abstract

**Background:** Kayano et al. estimated the potential number of COVID-19 cases and deaths in Japan from February 17 to November 30, 2021, in a counterfactual scenario where no vaccination was implemented. Their model predicted 63.3 million cases and 364,000 deaths, with a claimed 95% confidence interval of less than 1% of the estimated values.

**Objective:** To validate the transmission model used by Kayano et al. by simulating infection counts in Japan during 2020 and assessing the impact of errors in the reproduction number of the Delta variant on infection count estimates in the counterfactual scenario.

**Methods:** We replicated the model used by Kayano et al. to simulate infection dynamics in Japan during 2020. We evaluated the model’s performance by comparing the simulated infection surges with actual data. Additionally, we analyzed the sensitivity of infection count estimates to ±10% errors in the reproduction number of the Delta variant, corresponding to the 95% confidence interval reported in the cited study.

**Results:** The model successfully reproduced the first infection surge in early 2020, but it failed to replicate the second and third infection surges in the summer and winter of 2020. Sensitivity analysis revealed that a ±10% error in the reproduction number led to an estimated infection count error range of −25% to +42%.

**Conclusion:** The results suggest potential limitations in the validity of the counterfactual simulation results presented by Kayano et al., particularly regarding the accuracy of infection count estimates and the confidence interval.

## Introduction

A paper published in October 2023 [1] claimed that the cumulative numbers of COVID-19 infections and deaths in Japan from February 17 to November 30, 2021, are estimated to be 63.3 million (95% confidence interval [CI] 63.2–63.6) and 364 thousand (95% CI 363–366) respectively in the absence of vaccination. In contrast, the actual reported figures were 1.2 million infections and 10 thousand deaths, suggesting that the vaccination program reduced mortality by more than 97% in Japan. This study also claims that cases and deaths could have been reduced by 54% and 48% respectively had the vaccination been implemented 14 days earlier.

The simulation results of the study mentioned above gained significant attention and were widely reported by many mainstream media outlets in Japan [2,3], partially due to the fame of the corresponding author, who played a leading role as an advisor to the Japanese government during the COVID-19 pandemic. While some medical professionals cited the paper to support the efficacy of Japan’s COVID-19 vaccination efforts, others have expressed skepticism [4-6], questioning the validity of the estimated figures given their large scale.

In the above study, counterfactual simulations of COVID-19 cases and deaths are based on an extended version of the SIR (susceptible, infected, recovered) model. The effective reproduction number, which is interpreted as the average number of secondary cases in age group *a* generated by a single primary case in age group *b* at calendar time *t*, is given by

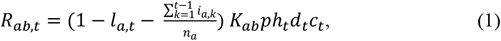

where *l*_*a*,*t*_ denotes the immune fraction in age group *aa* at calendar time (day) *t*, which is based on the estimated efficacy of vaccines, while 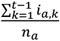 represents the cumulative number of previous infections. *K*_*ab*_ is a next-generation matrix given by a product of relative susceptibility of age group *aa* and the contact matrix between age groups *a* and *b. p* denotes the scaling parameter and *h*_*t*_ expresses the human mobility at time *t. d*_*t*_ represents the increase in transmissibility of the Delta variant compared with earlier variants and *c*_*t*_ expresses the influence of consecutive holidays at time *t*. The left bracketed term in equation (1), given by

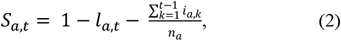

represents the fraction of susceptible populations in age group *a* at time *t*, assuming that none of those previously infected are susceptible.

The number of infections *i*_*a*,*t*_ with SARS-CoV-2 in age group *a* at time *t* is given by

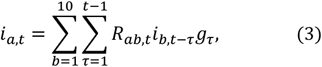

where *g*_*τ*_ indicates the probability density function of the generation interval, assumed to follow a Weibull distribution with a mean of 4.8 days and SD of 2.2 days. This modeling, however, does not reflect the real world, for the viral load of SARS-CoV-2 in an infected individual rises up soon after infection [7]. The confidence interval of infection count is calculated by fluctuating the number of infections with a Poisson distribution and applying the maximum likelihood estimation to the scaling parameters related to *p, h*_*t*_, *d*_*t*_, and *c*_*t*_.

Kayano et al. [1] changed *d*_*t*_ in equation (1) to reflect the increase of infectivity in the Delta variant. They refer to three previous studies for estimation of maximum *d*_*t*_ [8-10], while only one of them can be the basis of their assumption that the Delta variant is 1.5 times more infectious than the Alpha variant [9], where the Alpha variant is estimated to be 1.29 times more infectious (CI: 1.24-1.33) and the Delta variant is estimated to be 1.97 times more infectious (CI: 1.76-2.17) than the original Wuhan strain. Kayano et al., however, assume that *d*_*t*_ takes a fixed value with a probability of 100% and do not take into account the fluctuation in the estimation of *d*_*t*_ when they calculate the confidence interval of infection cases.

Kakeya et al. [11] compared the simulation results by Kayano et al. with the actual statistics, including prefectural data where vaccination programs were administered with varying time schedules to reassess the credibility of the modell. They found that the counterfactual scenario proposed by Kayano et al. does not account for the prefectural differences in infection rates resulting from different vaccination timings. This way of validation, however, is indirect and is not sufficient enough to completely refute the study by Kayano et al.

The authors repeatedly requested the corresponding author of the paper by Kayano et al. to share the source code in order to reproduce the results. Although he has consistently refused to provide the source code of the simulation, he has shared the parameters used in their simulation, namely, *h*_*t*_, *d*_*t*_, and *c*_*t*_.

This paper takes a more direct approach to challenge Kayano et al. by using their model and their parameters to simulate infections in Japan during 2020, in order to confirm whether their model can accurately predict the infection counts. Additionally, we analyzed how errors in the reproduction number affect the infection count in the counterfactual scenario where no vaccination was administered, in order to assess the reliability of their predictions.

## Methods

In this paper, a simplified transmission model given by

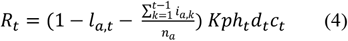

was used, since *K*_*ab*_ was only vaguely provided as a colormap. *K*_*ab*_ plays an important role when simulating death counts, since the death rate heavily depends on the age of the infected. However, it does not have a large influence when simulating case counts, which is the focus in this study.

The number of infections in this paper was given by

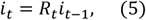

which is also a simplified model, for equation (3) is not reliable enough for the reason mentioned above and using equation (5) in place of (3) does not significantly affect the behavior of the simulation.

The value of *i*_*t*_ can be extremely small, potentially less than 1, depending on the experimental conditions, which can influence the subsequent result. To mitigate this, a minimum value of 0.1 was applied to *i*_*t*_. To reflect the delay in infection and report after infection, a five-day delay was added when comparing the simulation results with the real-world data [12].

The value of *Kp* in equation (4) was estimated to adjust the total number of infection cases in the counterfactual simulation. First, based on the estimated value of *Kp*, the infection counts in Japan during 2020 were simulated using the above model. Since the strain predominant during 2020 is reported to have been less infectious than the Alpha variant, which was predominant in early 2021, the value of *d*_*t*_ was reduced to account for that factor. Since *c*_*t*_ in [1] was almost always close to 1, it was fixed at 1 during the simulation of infection in 2020. Second, the variance in the total infection cases given by the above transmission model was calculated when the value of *d*_*t*_ changes within the confidence interval of the cited paper [9].

## Results

Figure 1 shows the result of the counterfactual simulation when *Kp* = 3.5127 and the parameters *h*_*t*_, *d*_*t*_, and *c*_*t*_ used by Kayano et al. are applied. Since this is a counterfactual simulation, the effect of immunization with vaccines is not considered. *Kp* was adjusted so that the total count of infections by Kayano et al. is reproduced. A general shape of the infection surge is reproduced successfully, though the height of the first small surge from April to May is smaller in the simulation than in the real-world data (7-day average). The simulation results heavily depend on the initial value. When the initial date is moved to February 28, 2021 and *Kp* = 3.4845, the height of the first surge comes close to the real-world data, while the total count of infections by Kayano et al. is also reproduced.

**Figure 1.**
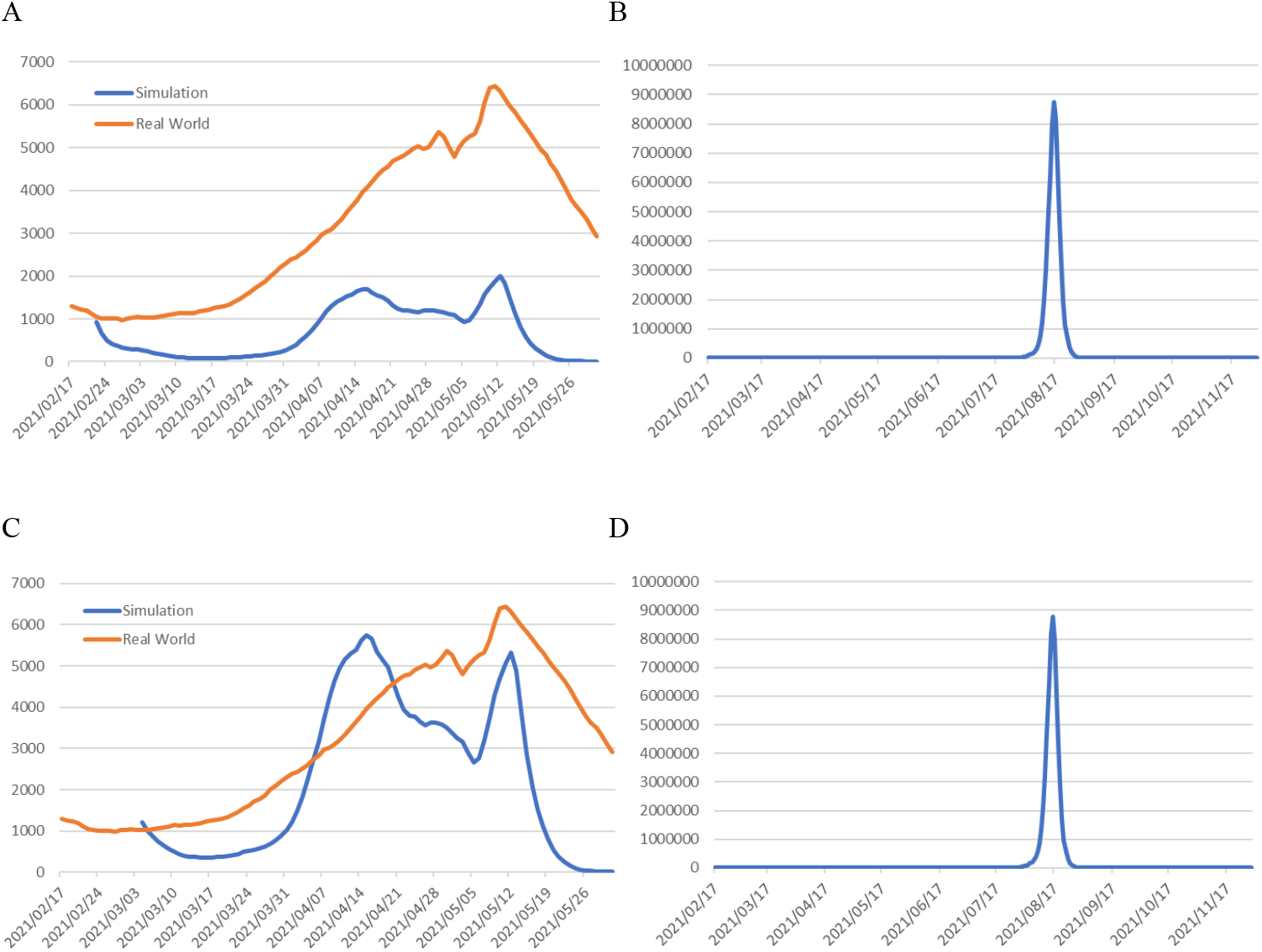
Infection counts during 2021 from a counterfactual simulation in the absence of the vaccination program when the parameters *h*_*t*_, *d*_*t*_, and *c*_*t*_ used by Kayano et al. are applied. (A): *Kp* = 3.5127, February-May, starting February 17; (B): *Kp* = 3.5127, February-November, starting February 17; (C): *Kp* = 3.4845, February-May, starting February 28; (D): *Kp* = 3.4845, February-November, starting February 28.

Figure 2 shows the simulation result for 2020 based on the transmission model given by equations (4) and (5) and the human mobility in 2020, where *Kp* = 3.0. As the figure shows, the first infection surge in early 20 20 is successfully reproduced, though the timing and the height of peak do not exactly match, while the second and third surges in the summer and winter of 2020 observed in the real-world data are not reproduced.

**Figure 2.**
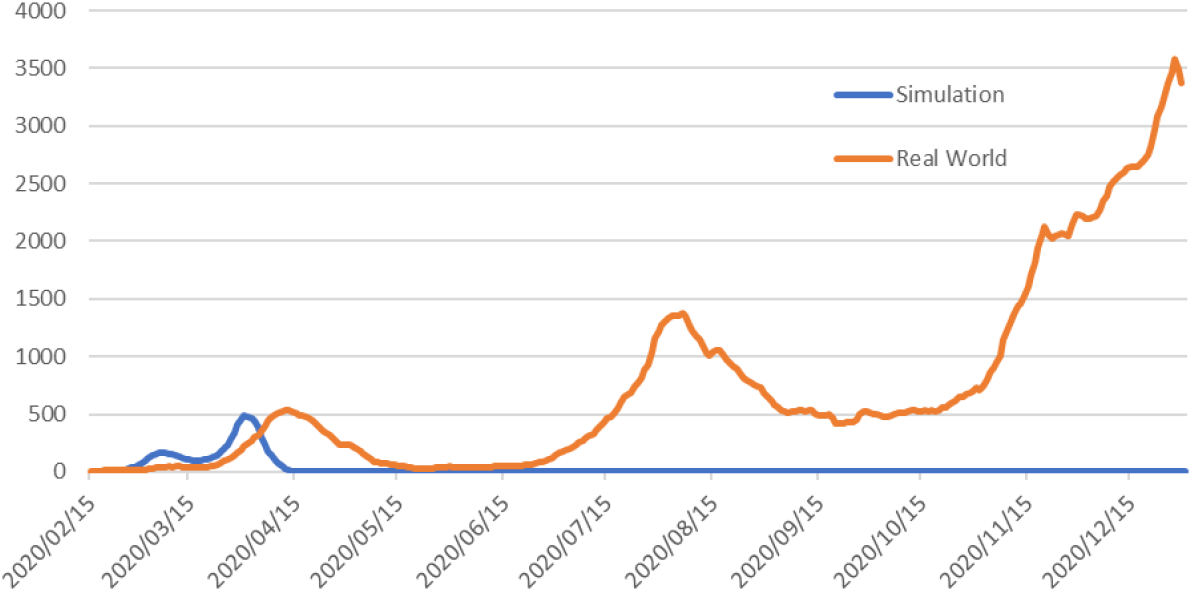
Infection counts from a simulation based on the transmission models given by equations (4) and (5), and the human mobility in 2020, where *Kp* = 3.0.

Figure 3 shows the results of the counterfactual simulation when max *d*_*t*_was fluctuated between −10% and +10%, reflecting the confidence interval of the paper cited by Kayano et al. [9]. As the figure shows, the height of the peak and the total count of infections drastically changes. It is also noteworthy that an unexpected bump appears in the end of 2021 when max *d*_*t*_ is 0.9 times as large as that of the standard simulation.

**Figure 3.**
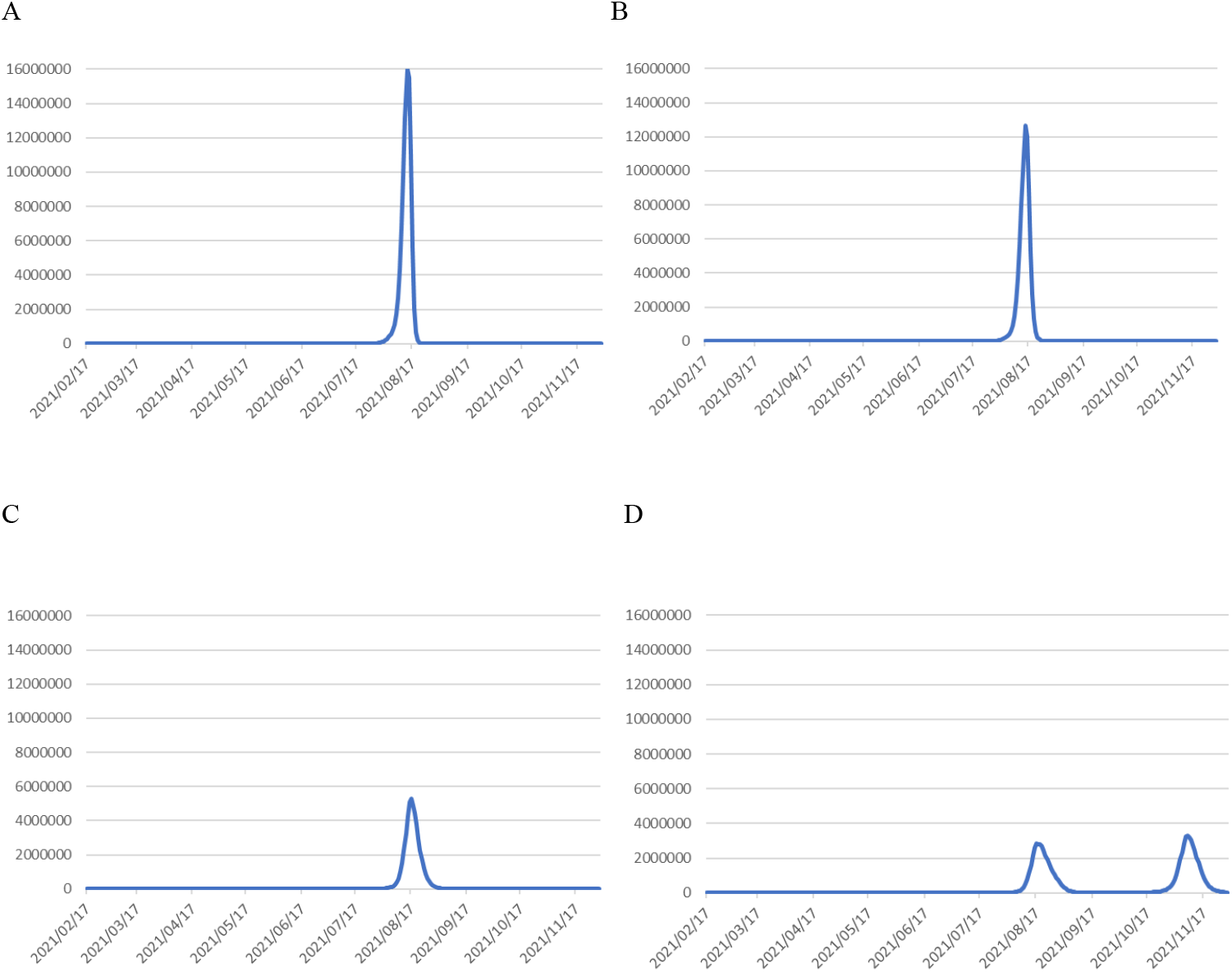
Results of the counterfactual simulation when max *d*_*t*_ is (A) +10%, (B) +5%, (C) −5%, and (D) −10% compared to the condition in Figure 1.

Figure 4 shows the change in the total counts of infections under different values of max *d*_*t*_. As the figure shows, the total count of infections ranges between −25% and +42%, which is extremely larger than the estimate by Kayano et al.

**Figure 4.**
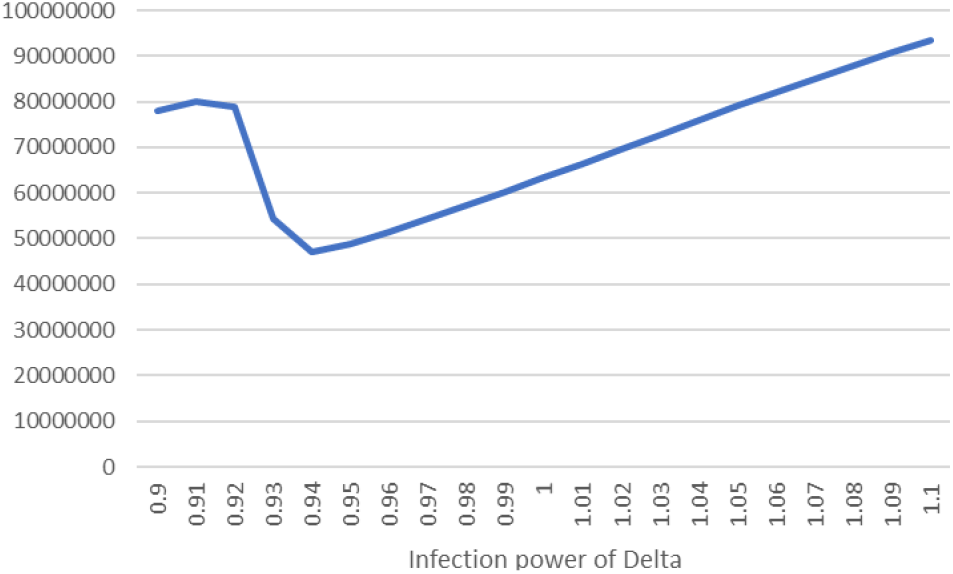
Change in the total count of infections under different max *d*_*t*_. The infection power is set to unity when *Kp* = 3.5127.

## Discussion

The results obtained in this paper cast serious doubt on the credibility of the simulation results by Kayano et al. Kayano et al. do not integrate seasonal factors into their model. It is widely known that surges in COVID-19 infections have occurred seasonally, with large peaks in winter and smaller peaks in summer [13], which is considered to be caused by reduced ventilation due to seasonal room cooling and heating. The failure to reproduce the surges in the summer and winter of 2020 is likely due to the lack of seasonal factors in their transmission model. It is known that SARS-CoV-2 is an airborne virus [14], and a lack of ventilation increases the risk of transmission significantly. The inability to reproduce the surges in the summer and winter of 2020 can be explained by the omission of seasonal factors in the transmission model by Kayano et al.

The simulation results with different *d*_*t*_ values highlight the non-robust nature of the transmission model, which is easily explained by its exponential nature. The second bump that appeared in one of the simulations can be explained by the coincidental overlap of surges in human mobility and in the number of infections. In the real world, human mobility depends on the temporal number of infections and should be expressed as a function of the number of infections. Using the human mobility from the factual scenario in the counterfactual simulation is not a valid way to estimate the number of infections in a counterfactual scenario, as evidenced by the emergence of the unexpected bump in one of the simulations.

It is true that the simulation in this paper is based on a simplified model as the detailed calculation method used by Kayano et al. is unknown owing to inability to access the original source code. However, it is strongly suggested that the credibility of the model by Kayano et al. is still an open question, as neither cross-validation of the model by the authors themselves nor follow-up experiments by peer researchers have ever been achieved so far. A simulation without cross-validation or any other validation with real-world data does not meet the standards of academic research. Likewise, it is inadequate to derive a confidence interval without taking into account the potential errors in parameters that can significantly affect the simulation result.

It is often pointed out that publications in medical journals tend to place too much emphasis on the implications for the medical community rather than on scientific rationality and integrity [15]. As concerns about the potential harms of COVID-19 mRNA vaccinations are being raised more often and openly [16], overestimation of the benefits of vaccination to justify its use in risk-benefit analyses for political purposes should never be accepted in order to maintain objectivity and integrity of science. Rigorous scientific studies to estimate the merits of vaccination are needed to establish sound and reliable future public health policy.

## Data availability statement

All the data used for the study are available from the corresponding author upon request.

## Author contributions

HK performed the simulations, and YU supervised the study. HK and YU co-authored the manuscript.

## Acknowledgements

The authors thank Prof. Hiroshi Nishiura for partially providing the simulation parameters used by his research group. The authors also thank Profs. Yasushi Iwamoto and Takashi Nakamura for their comments and suggestions on this study.

